# 2008 financial crisis vs 2020 economic fallout: How COVID-19 might influence fertility treatment and live births

**DOI:** 10.1101/2020.10.18.20214650

**Authors:** Piotr S. Gromski, Andrew D.A.C Smith, Deborah A Lawlor, Fady I. Sharara, Scott M Nelson

## Abstract

**Background:** The economic and reproductive medicine response to the COVID-19 pandemic in the United States has reduced the affordability and accessibility of fertility care. We sought to determine the impact of the 2008 financial recession and the COVID-19 recession on fertility treatments and cumulative live-births.

**Methods:** We examined annual US natality, CDC IVF cycle activity and live birth data from 1999 to 2018 encompassing 3,286,349 treatment cycles, to estimate the age-stratified reduction in IVF cycles undertaken after the 2008 financial recession, with forward quantitative modelling of IVF cycle activity and cumulative live-births for 2020 to 2023.

**Results:** The financial recession of 2008 caused a four-year plateau in fertility treatments with a predicted 53,026 (95% CI 49,581 to 56,471) fewer IVF cycles and 16,872 (95% CI 16,713 to 17,031) fewer live births. A similar scale of economic recession would cause 67,386 (95% CI: 61,686 to 73,086) fewer IVF cycles between 2020 and 2023, with women younger than 35 years overall undertaking 22,504 (95% CI 14,320 to 30,690) fewer cycles, as compared to 4,445 (95% CI 3,144 to 5749) fewer cycles in women over the age of 40 years. This equates to overall 25,143 (95% CI: 22,408 to 27,877) fewer predicted live-births from IVF, of which only 490 (95% CI 381 to 601) are anticipated to occur in women over the age of 40 years.

**Conclusions:** The COVID-19 recession could have a profound impact on US IVF live-birth rates in young women, further aggravating pre-existing declines in total fertility rates.

**Trial registration number:** not applicable

## Introduction

Affordability and availability of treatment are two of the most important factors affecting a couple’s decision to pursue in vitro fertilization (IVF). The emergence of COVID-19 as a risk to public health, and the resulting economic impact, affects both the affordability and availability of treatment. On March 17, 2020, the American Society for Reproductive Medicine (ASRM) recommended suspension of initiation of IVF treatments^1^. At least 85% of IVF clinics followed the recommendations and shut down provision of routine care. On April 24, the ASRM Task Force recommended “gradually and judiciously resuming the delivery of reproductive care”, with sequential updates reiterating this position given the dynamic situation.

Prior to the emergence of SARS-CoV-2, the US unemployment rate was at a historic low of 3.7% (5.7 million), and the economy at a peak^2^. With the onset of the COVID-19 pandemic, there was a dramatic rise in temporary unemployment peaking at 23.1 million in April 2020. However, despite a gradual decline to 17.8 million by June 2020, this has been accompanied by a rise in the number of permanent job losses to 2.9 million (Bureau of Labour Statistics July 2, 2020)^3^. In the developed world there is a pro-cyclical relationship between economic growth and fertility, and in times of economic recession the birth rate drops^4^. This was most recently observed after the 2008 financial crisis, where US birth rates declined and an estimated 2.3 million fewer births occurred between 2008 and 2013^5^. Clearly, economic hardship affects the affordability of having children and the decision to postpone is a potentially viable option for young women who have a longer fertility horizon^4^. For couples considering IVF, postponement incurs the penalty of an age-related declines in success^6^, while economic hardship additionally affects the affordability of treatment.

Reassuringly the COVID-19 related temporary closure of IVF units and accompanying treatment delays is anticipated to have limited impact on live-birth rates^7^. However, the impact of COVID-19 economic recession on IVF live births is unknown. Indeed, we are not aware of any prior publications regarding the impact of economic recessions on the use of IVF. COVID-19 additionally carries a direct health effect, fear of transmission, and fear of the unknown regarding pregnancy during a pandemic. The aim of this study is to examine the effect of the 2008 recession on IVF cycles and predict what impact the COVID-19 related economic recession and additional impact of clinic closures will have on the number of IVF cycles and live-births in the US.

## Methods

### Data sources

The Fertility Clinic Success Rate and Certification Act of 1992 requires that all assisted reproductive technology (ART) cycles performed in the United States are reported to the Centers for Disease Control and Prevention (CDC). Fertility clinics submit data to the CDC through the National ART Surveillance System (NASS) reporting system or an approved alternative compliant with federal reporting requirements. The CDC conducts data validation through yearly audits and site visits. The CDC has published Assisted Reproductive Technology Fertility Clinic Success Rates Reports detailing activity levels at an individual clinic level annually since 1997.

The Society for Assisted Reproductive Technology (SART), an organization of ART providers affiliated with the American Society for Reproductive Medicine (ASRM), has been collecting data and publishing annual reports of pregnancy success rates for fertility clinics in the United States and Canada since 1989. In 2017, of all the ART clinics reporting data to CDC, 82% were SART members.

Population comparison data was obtained from the 1999 to 2018 U.S. Natality files (Birth Cohort dataset) compiled annually by the CDC’s National Center for Health Statistics (NCHS). The NCHS provides information on 99% of all registered births each year in the United States.

### Definitions

We defined ART procedures as per the CDC, as all treatments or procedures that include the handling of human eggs or embryos to help a woman become pregnant. We defined a cycle of IVF as commencement of ovarian stimulation, or monitoring, with the intent of having an oocyte retrieval. This definition has been used by SART since 2014 and the CDC since 2017. This definition incorporates ovarian stimulation cycles which are cancelled, pre-implantation genetic testing is undertaken, or all embryos are frozen. For SART data before 2014 and CDC data from 1999 to 2016 a cycle was defined as “Fresh Embryos from Nondonor Eggs” (Table S1). Due to the different cycle definition used by the CDC for 2014 to 2016 (Table S2), an age-stratified multiplication value was derived using the aligned 2017 and 2018 SART and CDC records (Table S3), and applied to the CDC data for 2014 to 2016. The results from data transformation using the multiplication factor are presented in Table S4.

We defined a live-birth as delivery of one or more infants with any signs of life. The cumulative live-birth rate was defined as the probability of a live-birth from an ovarian stimulation encompassing all subsequent fresh and frozen embryo transfers from that stimulation. The total number of infants born allowing for multiples was determined from the annual Natality files and CDC ART reports (Table S5).

Age is the most important predictor of live birth following IVF treatment. We therefore stratified our analyses by age, using age categories consistent with the CDC and SART: less than 35 years, 35-37 years, 38-40 years, 41-42 years, more than 42 years.

### Quantitative modelling on cycle activity

A quantitative prediction model was built using CDC data from 1999 to 2008, with four years onward prediction for 2009 to 2012. The predicted clinical activity was compared to the observed clinical activity after the 2008 financial recession and the percentage reduction in activity for each age category calculated (Table S7, S8).

A similar quantitative model was built to predict age-stratified cycle starts for 2020 to 2023, using baseline data from 2014 to 2018. We then applied the same percentage reduction in activity observed after 2008 to the period 2020 to 2023, for each age category (Table S9).

This assumes that the impact of COVID-19 on cycle activity equates to a reduction like that observed after the 2008 financial recession. As sensitivity analyses, we modelled a less severe economic decline for the impact of COVID-19, by reducing the percentage reduction in activity by a factor of 0.5, and a more severe decline by increasing it by a factor of 0.5.

### Impact of reduction in activity on live births

For the period 2009 to 2012 we multiplied the predicted cycle activity by the age-stratified cumulative live-birth rates as reported by the CDC in each of the respective annual ART Success Rates Reports. This predicted number of live-births for 2009 to 2012 was then compared to the observed live-births in the CDC ART annual reports.

For the period 2020 to 2023 we multiplied the different levels of predicted cycle activity (no recession, recession equivalent to 2008, less and more severe recession) by the age-stratified cumulative live birth rates reported in the most recent annual CDC 2018 report^3^. This details the cumulative live-birth rates from 135,673 stimulation cycles undertaken by the 448 clinics in the USA that were commenced between January 1, 2017 and December 31, 2017, with inclusion of all embryo transfers that occurred within 12 months, and live-birth follow-up to October 2018^8^.

In addition to our main analyses (modelling the predicted impact of the economic recession due to COVID-19), we modelled the predicted impact of the two months closure of ART clinics. We previously showed that a shutdown of IVF treatment centers would result in a reduction in live-birth rate, and this reduction would differ with age^9^. In the current study, we calculated the reduction caused by a two-month shutdown and applied this to the live-birth rate for each age strata in 2020 only, assuming that a shutdown would only occur in 2020.

### Statistical analysis

The R 4.0.0 software environment was used for data analysis. “Forecast” package was used to perform auto regressive integrated moving average (ARIMA) prediction^10^.

## Results

Figure 1 demonstrates the increase in IVF treatment provision over the last two decades, the increasing number of ART infants and their increasing contribution to all US births. The financial recession of 2008 was associated with the beginning of a decline in all US births, which has continued to present day and is predicted to continue (Figure 1, Table S6). In contrast, for ART there was evidence of a four-year plateau before recommencing an increase (Figure 1). During this plateau an estimated 53,026 (95% CI, 49,581 to 56,471) fewer IVF cycles were undertaken, increasing from 5,625 (95% CI, 5,467 to 5,788) fewer cycles in 2009 to 21,321 (95% CI, 19,589 to 23,053) by 2012, assuming similar underlying rates of growth prior to 2008 would have continued (Table S7).

**Figure 1:**
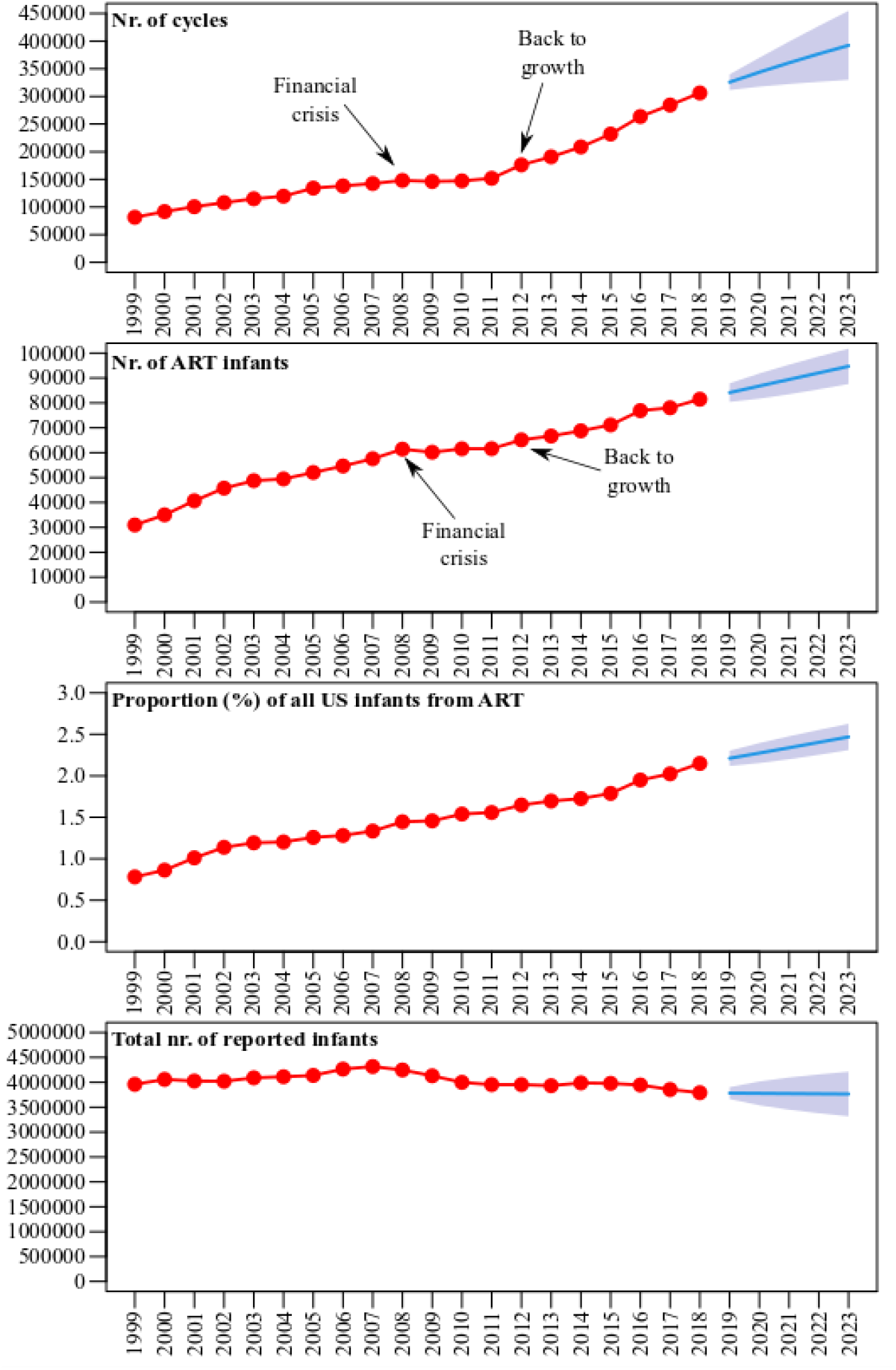
ART cycles, ART infants and all US births from 1999 to 2018 with 5 years prediction. The observed (red points) total number of ART treatments undertaken in the US (top panel), the total number of ART infants born per annum (second panel), and the proportion of ART infants (third panel) as a percentage of the total number of infants born within the US for 1999 to 2018 (bottom panel), with onward prediction for 2019 to 2023 (solid blue line with 95% confidence intervals). Actual values for number of all and ART infants for 1999 to 2018 are provided in Supplemental Table S2 with predictions provided in Table S6.

There was strong evidence of an age-specific reduction over the ensuing four years; women aged less than 35 years undertook 5.2% (95% CI, 1.1 to 8.9) fewer IVF cycles in 2009 with a further reduction to 15.8% (95% CI, 1.0 to 27.8) fewer cycles in 2012, as compared to women aged more than 40 years where the reduction was 2.9% (95% CI, 0.9 to 6.3) in 2009 and 6.5% (95% CI −9.0% to 18.1%) in 2012 (Figure 2, Table S7). This estimated reduction in cycle activity between 2009 and 2012 equates to 16,872 (95% CI, 16,713 to 17,031) predicted fewer live births, with these predominantly being derived from younger women due to their higher success rates (Table S9).

**Figure 2:**
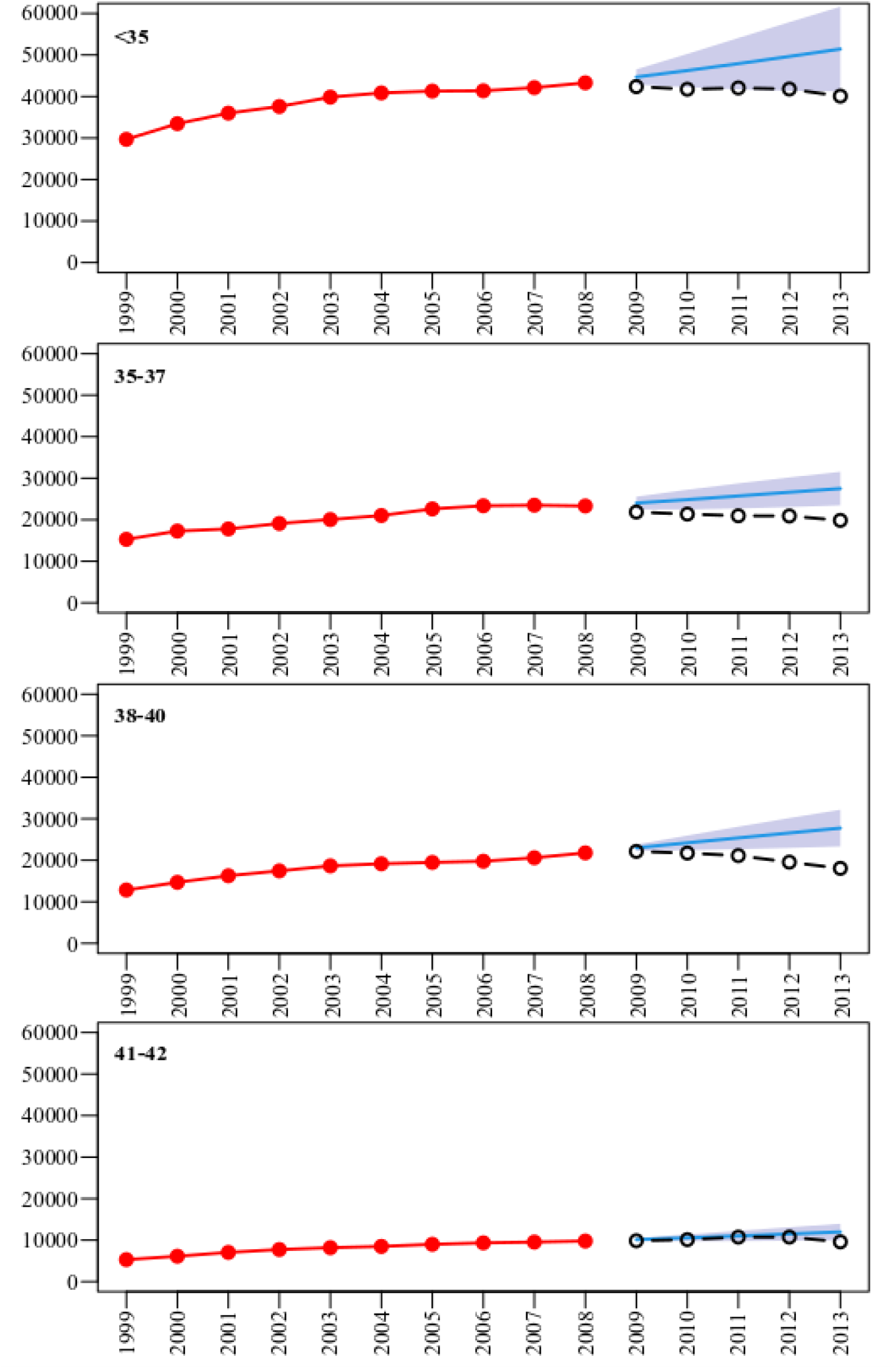
Observed and projected IVF cycle activity around 2008 financial crisis. The observed age-stratified number of cycles between 1999 and 2008 (red solid points), was used to predict number of cycles for 2009 to 2013 (solid blue line with 95% confidence intervals), as compared to the observed number of cycles (green open circles).

Given the underlying growth of IVF treatment cycles we estimate that 137,760 (95% CI, 13,486 to 149,034) IVF treatment cycles would have been initiated in 2020, increasing to 151,690 (95% CI, 123,321 to 180,059) in 2023 (Table 1). Estimation of the effect of COVID-19 economic recession on IVF activity would predict that 67,386 (95% CI, 61,686 to 73,086) fewer IVF cycles will occur over this time frame, mostly from women younger than 35 years (Figure 3, Table 1).

**Table 1.** Estimated changes in number of IVF fresh nondonor cycles over the period 2020-2023 with implication of economic crisis triggered by COVID-19 by age of patients.

| Age group<br>(years) | Without COVID-19 impact<br>Nr. of cycles (95% CI) |  | Percentage<br>reduction in cycles | With COVID-19 impact<br>Nr. of cycles (95% CI) |  | Difference (95% CI) |  |
| --- | --- | --- | --- | --- | --- | --- | --- |
| Year: 2020 |  |  |  |  |  |  |  |
| < 35 | 51,170 | (43,798~58,542) | -5.16% | 48,530 | (41,538~55,522) | -2,640 | (-3,020~-2,260) |
| 35-37 | 31,358 | (28,389~34,327) | -9.00% | 28,536 | (25,834~31,238) | -2822 | (-3,089~-2,555) |
| 38-40 | 30,385 | (27,296~33,474) | -3.76% | 29,243 | (26,270~32,216) | -1142 | (-1,259~-1,026) |
| 41-42 | 14,924 | (13,234~16,615) | -2.85% | 14,499 | (12,857~16,141) | -425 | (-474~-377) |
| >42 | 11,923 | (8,691~15,155) | -2.85% | 11,583 | (8,443~14,723) | -340 | (-432~-248) |
| Total | 139,760 | (130,486~149,034) | -5.27% | 132,391 | (123,583~141,199) | -7,369 | (-7,835~-6,903) |
| Year: 2021 |  |  |  |  |  |  |  |
| < 35 | 51,835 | (36,545~67,126) | -9.75% | 46,781 | (32,981~60,581) | -5,054 | (-6,545~-3,564) |
| 35-37 | 32,771 | (27,096~38,446) | -14.06% | 28,163 | (23,286~33,040) | -4,608 | (-5,406~-3,810) |
| 38-40 | 31,704 | (25,583~37,825) | -10.20% | 28,470 | (22,973~33,967) | -3,234 | (-3,858~-2,610) |
| 41-42 | 15,469 | (11,670~19,268) | -4.11% | 14,833 | (11,190~18,476) | -636 | (-792~-480) |
| >42 | 11,935 | (8,553~15,318) | -4.11% | 11,444 | (8,201~14,687) | -491 | (-631~-352) |
| Total | 143,714 | (125,566~161,862) | -9.76% | 129,691 | (98,633~160,752) | -14,023 | (-15,794~-12,252) |
| Year: 2022 |  |  |  |  |  |  |  |
| < 35 | 52,499 | (32,207~72,792) | -12.21% | 46,089 | (28,274~63,904) | -6410 | (-8,888~-3,933) |
| 35-37 | 34,222 | (27,014~41,431) | -18.56% | 27,870 | (21,999~33,741) | -6352 | (-7,690~-5,015) |
| 38-40 | 33,041 | (25,126~40,956) | -16.81% | 27,487 | (20,902~34,072) | -5554 | (-6,884~-4,224) |
| 41-42 | 16,034 | (10,707~21,362) | -2.52% | 15,630 | (10,437~20,823) | -404 | (-539~-270) |
| >42 | 11,913 | (8,507~15,318) | -2.52% | 11,613 | (8,293~14,933) | -300 | (-385~-214) |
| Total | 147,709 | (123,910~171,508) | 12.88% | 128,689 | (107,876~149,502) | -19,020 | (-22,006~-16,034) |

| Year: 2023 |  |  |  |  |  |  |  |
| --- | --- | --- | --- | --- | --- | --- | --- |
| < 35 | 53,163 | (28,878~77,448) | -15.80% | 44,763 | (24,315~65,211) | -8,400 | (-12,237~-4,563) |
| 35-37 | 35,665 | (27,153~44,177) | -21.43% | 28,022 | (21,334~34,710) | -7,643 | (-9,467~-5,819) |
| 38-40 | 34,376 | (24,987~43,765) | -26.42% | 25,294 | (18,386~32,202) | -9,082 | (-11,563~-6,601) |
| 41-42 | 16,603 | (10,059~23,147) | -6.49% | 15,525 | (9,406~21,644) | -1,078 | (-1,503~-653) |
| >42 | 11,883 | (8,474~15,293) | -6.49% | 11,112 | (7,924~14,300) | -771 | (-993~-550) |
| **Total** | **151,690** | **(123,321~180,059)** | **-17.78%** | **124,716** | **(101,090~148,342)** | **-26,974** | **(-31,718~-22,230)** |

**Figure 3:**
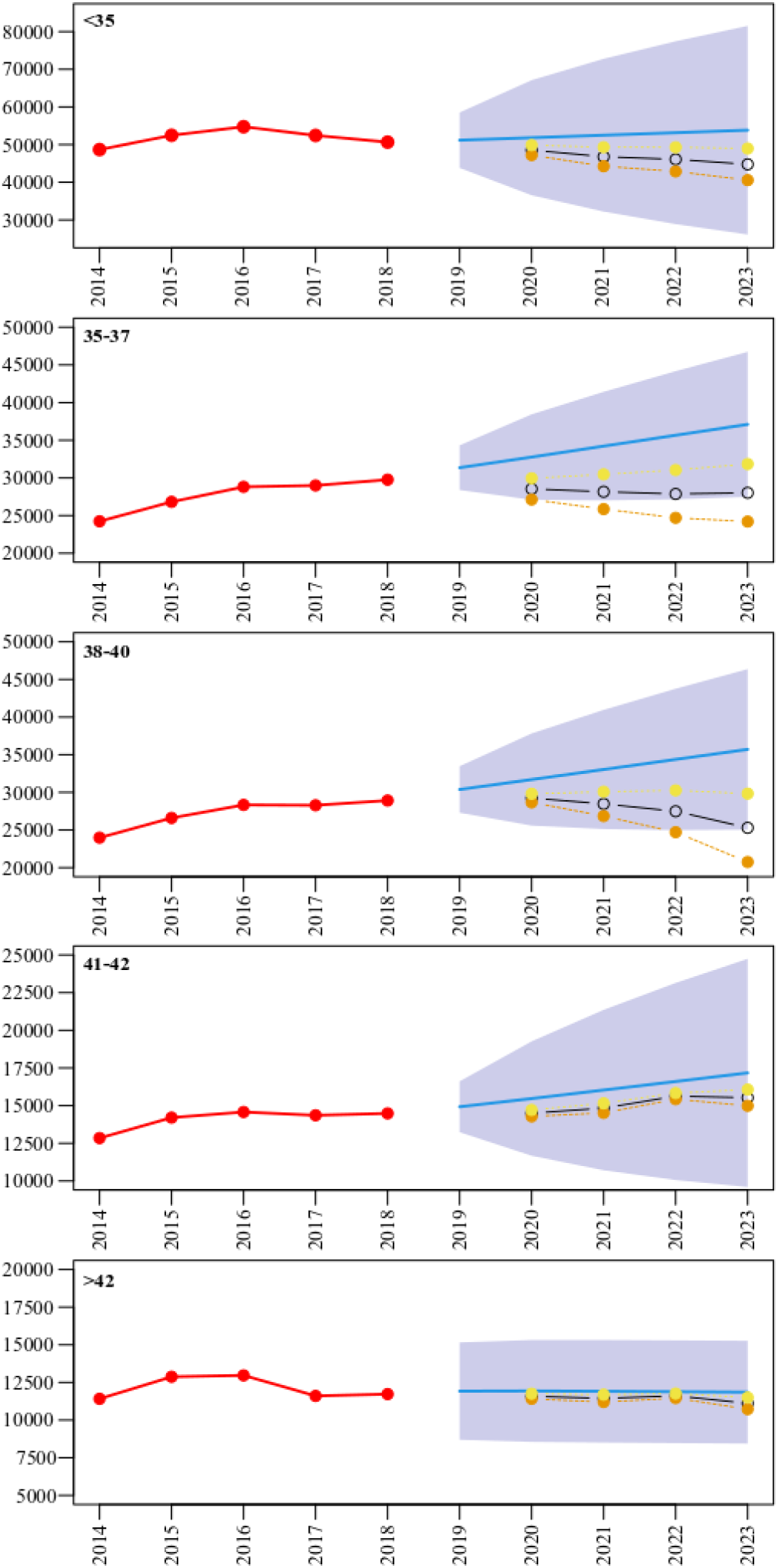
Observed and projected IVF cycle activity in response to COVID-19 financial crisis. The observed age-stratified number of cycles between 2013 and 2018 (red solid points), was used to predict number of cycles for 2019 to 2023 (solid blue line with 95% confidence intervals). The estimated shifts reflect the age strata specific percentage decline observed after the 2008 crisis (green line), with a 50% less severe impact (brown solid line) or 50% more severe impact (black line).

With the closure of the IVF units for 2 months, the minor increase in maternal age will be associated with a small reduction (−0.7% (95% CI, −1.0 to −0.3%)) on the overall cumulative live-birth rates for treatments initiated over the whole population in 2020 (Table S10 and S11). This delay combined with the reduction in clinical activity due to COVID-19, is predicted to result in 3,414 (95% CI, 3,193 to 3,636) fewer live-births in 2020 (Table 2). With the overall (combining the impact of the economic recession and clinic closures) estimated reduction in IVF activity for 2020 to 2023, we predict 25,143 (95% CI, 22,408 to 27,877) fewer live births (Table 2). Sensitivity analyses estimating the impact of less and more severe economic recession following COVID-19 on IVF cycles and live births are shown in Tables S12 to S15.

**Table 2.** Estimated changes in number of IVF live births over the period 2020-2023 with implication of economic crisis triggered by COVID-19 by age of patients.

| Age group (years) | Without COVID-19 impact |  | With COVID-19 impact |  | Difference (95% CI) |  |
| --- | --- | --- | --- | --- | --- | --- |
|  | Live births (95% CI) |  | Live births (95% CI) |  |  |  |
| Year: 2020 |  |  |  |  |  |  |
| < 35 | 26,404 | (22,593~30,214) | 24,884 | (21,293~28,475) | -1,520 | (-1,716~-1,324) |
| 35-37 | 11,759 | (10,632~12,886) | 10,487 | (9,482~11,493) | -1,272 | (-1,372~-1,172) |
| 38-40 | 7,140 | (6,399~7,882) | 6,705 | (6,009~7,401) | -435 | (-463~-408) |
| 41-42 | 1,761 | (1,547~1,975) | 1,611 | (1,414~1,807) | -150 | (-156~-145) |
| >42 | 405 | (289~522) | 369 | (262~475) | -37 | (-40~-34) |
| Total | 47,470 | (43,421~51,519) | 44,056 | (40,255~47,856) | -3,414 | (-3,636~-3,193) |
| Year: 2021 |  |  |  |  |  |  |
| < 35 | 26,747 | (18,854~34,640) | 24,139 | (17,016~31,262) | -2,608 | (-3,377~-1,838) |
| 35-37 | 12,289 | (10,153~14,425) | 10,561 | (8,726~12,397) | -1,728 | (-2,028~-1,428) |
| 38-40 | 7,450 | (6,003~8,897) | 6,690 | (5,391~7,990) | -760 | (-908~-612) |
| 41-42 | 1,825 | (1,370~2,281) | 1,750 | (1,313~2,187) | -75 | (-94~-56) |
| >42 | 406 | (284~527) | 389 | (273~506) | -17 | (-22~-12) |
| Total | 48,718 | (40,400~57,035) | 43,530 | (36,046~51,014) | -5,188 | (-6,027~-4,348) |
| Year: 2022 |  |  |  |  |  |  |
| < 35 | 27,089 | (16,616~37,563) | 23,782 | (14,587~32,976) | -3,308 | (-4,586~-2,029) |
| 35-37 | 12,833 | (10,123~15,543) | 10,451 | (8,244~12,658) | -2,382 | (-2,885~-1,879) |
| 38-40 | 7,765 | (5,897~9,632) | 6,459 | (4,906~8,013) | -1,305 | (-1,619~-991) |
| 41-42 | 1,892 | (1,258~2,526) | 1,844 | (1,226~2,463) | -48 | (-64~-32) |
| >42 | 405 | (283~527) | 395 | (276~514) | -10 | (-13~-7) |
| Total | 49,984 | (38,987~60,982) | 42,932 | (33,329~52,535) | -7,053 | (-8,462~-5,643) |
| Year: 2023 |  |  |  |  |  |  |
| < 35 | 27,432 | (14,899~39,965) | 23,098 | (12,545~33,651) | -4,334 | (-6,315~-2,354) |
| 35-37 | 13,374 | (10,176~16,573) | 10,508 | (7,995~13,021) | -2,866 | (-3,551~-2,181) |
| 38-40 | 8,078 | (5,865~10,291) | 5,944 | (4,316~7,572) | -2,134 | (-2,719~-1,550) |
| 41-42 | 1,959 | (1,182~2,736) | 1,832 | (1,105~2,559) | -127 | (-178~-77) |
| >42 | 404 | (282~526) | 378 | (263~492) | -26 | (-34~-18) |
| Total | 51,248 | (38,102~64,394) | 41,760 | (30,766~52,754) | -9,488 | (-11,664~-7,312) |

## Discussion

We demonstrate that the enduring growth of US fertility treatments temporarily halted after the 2008 financial crisis until 2012 and then resumed. Despite widespread economic adversity at this time, women over 40 years largely continued to pursue treatment as compared to younger women. With economic indicators suggesting an equivalent or even greater recession anticipated secondary to COVID-19, we estimate that the pandemic will result in 67,386 fewer IVF cycles being undertaken and 25,143 fewer live-births in the US over the next four years, equating to 12.7% fewer women having a baby from IVF, with the greatest reduction observed in women less than 40 years old. This reduction will be primarily driven by the anticipated economic recession, with clinic closures making only a small contribution. We acknowledge that these predicted decreases in IVF conceived live-births following the 2008 recession contribute <2% of the reduction in all live-births in the US at that time, and despite recent growth ART still constitutes <3% of US births. Thus, reductions in fertility treatments following the current COVID-19 related recession are not anticipated to have a major impact on population levels in the US or any other country. Nonetheless the now accepted right of couples to control their fertility, including through access to treatments, is likely to be importantly impacted.

For assisted conception which is predominantly self-funded by patients or insurers, the loss of employment or financial security may have been responsible for the plateau in the aftermath of the 2008 financial recession. That women older than 40 aged exhibited the lowest percentage reduction in clinical activity potentially reflects their greater appreciation of the age-related decline in both spontaneous fecundity^17^ and IVF success rates^6,18^. Furthermore, a decision to not pursue fertility treatments would have the greatest impact on older women as assisted reproductive technology births equate to 11.8% of all births in women over the age of 40, as compared to 4.4% in women aged 35-39 years, and 0.9% in women under 35 years^19^. Older women are also likely to be more financially secure than younger women.

The disruptive economic repercussions of COVID-19 continue to be elucidated with the use of high-frequency indicators of economic fluctuations, such as unemployment insurance claims, which breached 30 million in the first six weeks of the pandemic, implying a dramatic reduction in future employment and labor force participation^3,20^. At present the longer-term projections exceed the 2008 crisis, and if the recovery is muted it could take more than 5 years for the most affected sectors to return to 2019-level contributions to GDP. This backdrop of financial uncertainty is likely to translate into a relative reduction in fertility related treatment. We anticipate that like 2008, the greatest reduction in clinical activity will be in younger women, who will perceive that they may be able to wait and conceive naturally and / or are unable to afford treatment. Reductions in IVF in this age-group will however have the largest overall impact on birth rates due to the volume of activity and relative high success rates. Given the relatively small contribution of ART births to all US births the greatest threat to the population is from younger women deciding to postpone natural conception or decide against further children due to economic uncertainty (Figure 1 or Table S5). That this would occur at a time when US fertility rates are already substantially below replacement levels will have further profound impacts on population age structures^21^.

Contracting economies may also affect health by impeding adherence to preventive measures or adoption of unhealthy lifestyle characteristics. Economic downturns have been associated with increases rates of obesity^23^, reduce attempts at smoking cessation^24^, increased incidence of sexually transmitted infections^25^ all of which would further impede spontaneous and assisted conception. Perinatal outcomes may also be compromised as economic adversity has been associated with an increased risk of miscarriage^26^ and stillbirths^27^.

Our results show that economic recession erodes the accepted rights of couples to have their desired family. Amid an absence of public funding, and patchwork of state mandates for insurance provision, an economic recession will exacerbate unequal access to health care, especially for minorities. Lack of an infertility insurance mandate has previously been associated with an increased risk of triplets and high order multiples, preterm birth and low birth weight^29^, as patients may seek more affordable treatments and take higher risks to address involuntary childlessness. In countries where health-care provision is equally accessible irrespective of employment or insurance status, access to fertility treatment will be less problematic and declining birth rates may be less exacerbated by COVID-19.

We note several limitations of our study. We evaluated population health outcomes and economic trends and did not account for variations at regional or subnational levels, which may mask variations particularly as the percentage of children born via assisted conception varies from high levels in some states such as Massachusetts (4.8%) to low in Puerto Rico (0.2%)^30^. For reasons of data availability and definitions we utilised total treatment cycles to develop our growth estimates, thereby allowing for variations in clinical practice and contextualisation of the 2008 financial crisis over two decades. We are unaware of any other reasons for the observed plateau, with the resumption of growth in 2012 aligning with other improvements in macroeconomic indicators. We have assumed an equivalent economic challenge due to COVID-19 as observed in 2008, however, we have performed sensitivity analyses for both a less or more severe impact and the true estimate on cycles and live births is likely to lie between these two extremes. Our prediction methods assume that past activity is a reliable indicator of future activity; we discuss these assumptions and show our predictions are not sensitive to them in Supplementary Material. Lastly, our estimates of live-birth reflect current reported success rates for all treatments performed within 12 months of the initiated stimulation cycle as per CDC and SART, we acknowledge that additional frozen embryo transfers may occur beyond this time-frame resulting in some additional live-births that are unaccounted for. It is also possible that improvements in treatment, which we cannot predict, will cause a relative increase in future live-birth rates for some age groups.

We demonstrate the detrimental impact of the 2008 economic crisis on the uptake of fertility treatment, and that older women largely persisted in seeking assistance. We estimate that the COVID-19 related economic recession will be associated with about 25,143 fewer live births over the next four years, with the greatest reduction observed in women who are 35-40 year olds where ART related births constitute 4.4% of all US births.

## Data Availability

"Data sources; The Fertility Clinic Success Rate and Certification Act of 1992 requires that all assisted reproductive technology (ART) cycles performed in the United States are reported to the Centers for Disease Control and Prevention (CDC); Fertility clinics submit data to the CDC through the National ART Surveillance System (NASS) reporting system or an approved alternative compliant with federal reporting requirements; The CDC conducts data validation through yearly audits and site visits; The CDC has published Assisted Reproductive Technology Fertility Clinic Success Rates Reports detailing activity levels at an individual clinic level annually since 1997; The Society for Assisted Reproductive Technology (SART) an organization of ART providers affiliated with the American Society for Reproductive Medicine (ASRM) has been collecting data and publishing annual reports of pregnancy success rates for fertility clinics in the United States and Canada since 1989; In 2017 of all the ART clinics reporting data to CDC 82% were SART members.
Population comparison data was obtained from the 1999 to 2018 US Natality files (Birth Cohort dataset) compiled annually by the CDCs National Center for Health Statistics (NCHS); The NCHS provides information on 99% of all registered births each year in the United States"

https://www.cdc.gov/art/artdata/index.html

## Notes

### Competing Interest Statement

No competing interests: "No funding bodies had any role in the study design data collection and analysis decision to publish or preparation of the manuscript; SMN has participated in Advisory Boards and received speakers fees from Beckman Coulter; Ferring; Finox; Merck; MSD and Roche Diagnostics; DAL has received grant funding for other studies not related to this one from government charity and industry funders including Roche Diagnostics and Medtronic; DAL is a member of the Editorial Board for BMC Medicine; PSG and ADACS have no conflict of interest; FIS receive the speakers fees from Ferring."

### Clinical Trial

Using available statistical data. The Fertility Clinic Success Rate and Certification Act of 1992 requires that all assisted reproductive technology (ART) cycles performed in the United States are reported to the Centers for Disease Control and Prevention (CDC). Fertility clinics submit data to the CDC through the National ART Surveillance System (NASS) reporting system or an approved alternative compliant with federal reporting requirements.

### Funding Statement

This work was supported by the National Institute for Health Research Biomedical Centre at the University Hospitals Bristol NHS Foundation Trust and the University of Bristol (DAL, and SMN), a European Research Council grant (DevelopObese; 669545 to DAL), a British Heart Foundation Grant (AA/18/7/34219 to DAL) and a National Institute for Health Research Senior Investigator award (NF-0616-10102 to DAL). DAL work in a Unit that receives support from the University of Bristol and the MRC (MC_UU_00011/6). The views expressed in this publication are those of the author(s) and not necessarily those of the NHS, the National Institute for Health Research or the Department of Health and Social Care, or any other funders mentioned here.

### Author Declarations

University of Glasgow

