## Supplementary material for "2008 financial crisis vs 2020 economic fallout: How COVID-19 might influence fertility treatment and live births": SI

Content

1. Data

2. Cumulative Live birth modelling

3. Outcomes of COVID-19 with lower economic impact

4. Outcomes of COVID-19 with greater economic impact

### 1. Data

In this study we have employed two data sets: CDC and SART. From CDC data set summary of assisted reproductive technology (ART) success rates report in excel format were copied from: https://www.cdc.gov/art/artdata/index.html. Each of the report's cycles started and was carried out in different increasing number of clinics each year, and the outcomes of these cycles, during each calendar year where deposited in excel spreadsheets. At this point, from each of downloaded spreadsheets since 1999 we have extracted the number of cycles reported for fresh nondonor eggs strata by age of patients. For this purpose, as can be observed in **Table S1** we have used abbreviations as defined in description spreadsheet and collected the number of cycles strata by patient's age along with total number of cycles per each calendar year since 1999 till 2018. The total number of cycles was used in order to highlight the change in the number of cycles due to finical crisis and the amount of time to come back to steady growth.

The data from 1999-2013 were used with the purpose of simulation of potential growth of the number of cycles as observed over five years doubt that financial crisis in 2008 did not happen. The ARIMA model estimate *circa* five years increase of the number of cycles either in fresh nondonor eggs or total number of cycles. This numbers then are used to estimate percentage decrease/slow down in the fresh nondonor or total number of cycles.

**Table S1. ART success rates report since 1999 for fresh embryo nondonor eggs cycles by age of patients.**

| Year | Abbreviation | Description | Nr fresh nondonor cycles | Total nr ART treatments |
|---|---|---|---|---|
| 1999 | FshNDCycle1 | (Fresh Nondonor Eggs) Number of cycles <35 | 29,682 | 81,483 |
| | FshNDCycle2 | (Fresh Nondonor Eggs) Number of cycles 35-37 | 15,291 | |
| | FshNDCycle3 | (Fresh Nondonor Eggs) Number of cycles 38-40 | 12,848 | |
| | FshNDCycle4 | (Fresh Nondonor Eggs) Number of cycles 41-42 | 5,302 | |
| 2000 | FshNDCycle1 | (Fresh Nondonor Eggs) Number of cycles <35 | 33,453 | 91,779 |
| | FshNDCycle2 | (Fresh Nondonor Eggs) Number of cycles 35-37 | 17,284 | |
| | FshNDCycle3 | (Fresh Nondonor Eggs) Number of cycles 38-40 | 14,701 | |
| | FshNDCycle4 | (Fresh Nondonor Eggs) Number of cycles 41-42 | 6,118 | |
| 2001 | FshNDCycle1 | (Fresh Nondonor Eggs) Number of cycles <35 | 35,984 | 100,552 |
| | FshNDCycle2 | (Fresh Nondonor Eggs) Number of cycles 35-37 | 17,791 | |
| | FshNDCycle3 | (Fresh Nondonor Eggs) Number of cycles 38-40 | 16,283 | |
| | FshNDCycle4 | (Fresh Nondonor Eggs) Number of cycles 41-42 | 7,044 | |
| 2002 | FshNDCycle1 | (Fresh Nondonor Eggs) Number of cycles <35 | 37,591 | 107,927 |

| | | | | |
|---|---|---|---|---|
| | FshNDCycle2 | (Fresh Nondonor Eggs) Number of cycles 35-37 | 19,110 | |
| | FshNDCycle3 | (Fresh Nondonor Eggs) Number of cycles 38-40 | 17,454 | |
| | FshNDCycle4 | (Fresh Nondonor Eggs) Number of cycles 41-42 | 7,733 | |
| 2003 | FshNDCycle1 | (Fresh Nondonor Eggs) Number of cycles <35 | 39,852 | 114,963 |
| | FshNDCycle2 | (Fresh Nondonor Eggs) Number of cycles 35-37 | 20,056 | |
| | FshNDCycle3 | (Fresh Nondonor Eggs) Number of cycles 38-40 | 18,660 | |
| | FshNDCycle4 | (Fresh Nondonor Eggs) Number of cycles 41-42 | 8,185 | |
| 2004 | FshNDCycle1 | (Fresh Nondonor Eggs) Number of cycles <35 | 40,853 | 119,551 |
| | FshNDCycle2 | (Fresh Nondonor Eggs) Number of cycles 35-37 | 21,019 | |
| | FshNDCycle3 | (Fresh Nondonor Eggs) Number of cycles 38-40 | 19,174 | |
| | FshNDCycle4 | (Fresh Nondonor Eggs) Number of cycles 41-42 | 8,487 | |
| 2005 | FshNDCycle1 | (Fresh Nondonor Eggs) Number of cycles <35 | 41,301 | 134,260 |
| | FshNDCycle2 | (Fresh Nondonor Eggs) Number of cycles 35-37 | 22,622 | |
| | FshNDCycle3 | (Fresh Nondonor Eggs) Number of cycles 38-40 | 19,485 | |
| | FshNDCycle4 | (Fresh Nondonor Eggs) Number of cycles 41-42 | 8,997 | |
| 2006 | FshNDCycle1 | (Fresh Nondonor Eggs) Number of cycles <35 | 41,369 | 138,198 |
| | FshNDCycle2 | (Fresh Nondonor Eggs) Number of cycles 35-37 | 23,376 | |
| | FshNDCycle3 | (Fresh Nondonor Eggs) Number of cycles 38-40 | 19,775 | |
| | FshNDCycle4 | (Fresh Nondonor Eggs) Number of cycles 41-42 | 9,346 | |
| 2007 | FshNDCycle1 | (Fresh Nondonor Eggs) Number of cycles <35 | 42,127 | 142,435 |
| | FshNDCycle2 | (Fresh Nondonor Eggs) Number of cycles 35-37 | 23,504 | |
| | FshNDCycle3 | (Fresh Nondonor Eggs) Number of cycles 38-40 | 20,612 | |
| | FshNDCycle4 | (Fresh Nondonor Eggs) Number of cycles 41-42 | 9,535 | |
| | FshNDCycle5 | (Fresh Nondonor Eggs) Number of cycles 43-44 | 4,814 | |
| 2008 | FshNDCycle1 | (Fresh Nondonor Eggs) Number of cycles <35 | 43,296 | 148,055 |
| | FshNDCycle2 | (Fresh Nondonor Eggs) Number of cycles 35-37 | 23,326 | |
| | FshNDCycle3 | (Fresh Nondonor Eggs) Number of cycles 38-40 | 21,793 | |
| | FshNDCycle4 | (Fresh Nondonor Eggs) Number of cycles 41-42 | 9,783 | |
| | FshNDCycle5 | (Fresh Nondonor Eggs) Number of cycles 43-44 | 4,907 | |
| 2009 | FshNDCycle1 | (Fresh Nondonor Eggs) Number of cycles <35 | 42,384 | 146,244 |
| | FshNDCycle2 | (Fresh Nondonor Eggs) Number of cycles 35-37 | 21,860 | |
| | FshNDCycle3 | (Fresh Nondonor Eggs) Number of cycles 38-40 | 22,144 | |
| | FshNDCycle4 | (Fresh Nondonor Eggs) Number of cycles 41-42 | 9,845 | |
| | FshNDCycle5 | (Fresh Nondonor Eggs) Number of cycles 43-44 | 4,857 | |
| 2010 | FshNDCycle1 | (Fresh Nondonor Eggs) Number of cycles <35 | 41,741 | 147,260 |
| | FshNDCycle2 | (Fresh Nondonor Eggs) Number of cycles 35-37 | 21,366 | |
| | FshNDCycle3 | (Fresh Nondonor Eggs) Number of cycles 38-40 | 21,739 | |
| | FshNDCycle4 | (Fresh Nondonor Eggs) Number of cycles 41-42 | 10,120 | |
| | FshNDCycle5 | (Fresh Nondonor Eggs) Number of cycles 43-44 | 4,501 | |
| 2011 | FshNDCycle1 | (Fresh Nondonor Eggs) Number of cycles <35 | 42,059 | 151,923 |
| | FshNDCycle2 | (Fresh Nondonor Eggs) Number of cycles 35-37 | 20,963 | |
| | FshNDCycle3 | (Fresh Nondonor Eggs) Number of cycles 38-40 | 21,128 | |
| | FshNDCycle4 | (Fresh Nondonor Eggs) Number of cycles 41-42 | 10,733 | |
| | FshNDCycle5 | (Fresh Nondonor Eggs) Number of cycles 43-44 | 4,744 | |

| | | | | |
|---|---|---|---|---|
| | FshNDCycle6 | (Fresh Nondonor Eggs) Number of cycles >44 | 1,586 | |
| 2012 | FshNDCycle1 | (Fresh Nondonor Eggs) Number of cycles <35 | 41,798 | 176,247 |
| | FshNDCycle2 | (Fresh Nondonor Eggs) Number of cycles 35-37 | 20,920 | |
| | FshNDCycle3 | (Fresh Nondonor Eggs) Number of cycles 38-40 | 19,556 | |
| | FshNDCycle4 | (Fresh Nondonor Eggs) Number of cycles 41-42 | 10,740 | |
| | FshNDCycle5 | (Fresh Nondonor Eggs) Number of cycles 43-44 | 5,050 | |
| | FshNDCycle6 | (Fresh Nondonor Eggs) Number of cycles >44 | 1,601 | |
| 2013 | FshNDCycle1 | (Fresh Nondonor Eggs) Number of cycles <35 | 40,083 | 190,773 |
| | FshNDCycle2 | (Fresh Nondonor Eggs) Number of cycles 35-37 | 19,853 | |
| | FshNDCycle3 | (Fresh Nondonor Eggs) Number of cycles 38-40 | 18,061 | |
| | FshNDCycle4 | (Fresh Nondonor Eggs) Number of cycles 41-42 | 9,588 | |
| | FshNDCycle5 | (Fresh Nondonor Eggs) Number of cycles 43-44 | 4,823 | |
| | FshNDCycle6 | (Fresh Nondonor Eggs) Number of cycles >44 | 1,379 | |
| 2014 | FshNDCycle1 | (Fresh Nondonor Eggs) Number of cycles <35 | 39,573 | 208,604 |
| | FshNDCycle2 | (Fresh Nondonor Eggs) Number of cycles 35-37 | 19,376 | |
| | FshNDCycle3 | (Fresh Nondonor Eggs) Number of cycles 38-40 | 17,617 | |
| | FshNDCycle4 | (Fresh Nondonor Eggs) Number of cycles 41-42 | 9,114 | |
| | FshNDCycle5 | (Fresh Nondonor Eggs) Number of cycles 43-44 | 5,131 | |
| | FshNDCycle6 | (Fresh Nondonor Eggs) Number of cycles >44 | 2,051 | |
| 2015 | FshNDCycle1 | (Fresh Nondonor Eggs) Number of cycles <35 | 39,302 | 231,936 |
| | FshNDCycle2 | (Fresh Nondonor Eggs) Number of cycles 35-37 | 19,023 | |
| | FshNDCycle3 | (Fresh Nondonor Eggs) Number of cycles 38-40 | 17,191 | |
| | FshNDCycle4 | (Fresh Nondonor Eggs) Number of cycles 41-42 | 8,872 | |
| | FshNDCycle5 | (Fresh Nondonor Eggs) Number of cycles 43-44 | 4,940 | |
| | FshNDCycle6 | (Fresh Nondonor Eggs) Number of cycles >44 | 1,762 | |
| 2016 | FshNDCycle1 | Frsh emb Frsh nondnr egg <35 | 36,625 | 263,577 |
| | FshNDCycle2 | Frsh emb Frsh nondnr egg 35-37 | 18,278 | |
| | FshNDCycle3 | Frsh emb Frsh nondnr egg 38-40 | 16,109 | |
| | FshNDCycle4 | Frsh emb Frsh nondnr egg 41-42 | 8,264 | |
| | FshNDCycle5 | Frsh emb Frsh nondnr egg >42 | 6,961 | |
| 2017 | ND_NumIntentRet1 | Nondonor eggs, All patients <35 | 52,428 | 284,385 |
| | ND_NumIntentRet2 | Nondonor eggs, All patients 35-37 | 28,996 | |
| | ND_NumIntentRet3 | Nondonor eggs, All patients 38-40 | 28,287 | |
| | ND_NumIntentRet4 | Nondonor eggs, All patients 41-42 | 14,358 | |
| | ND_NumIntentRet5 | Nondonor eggs, All patients ≥43 | 11,604 | |
| 2018 | ND_NumIntentRet1 | Nondonor eggs, All patients <35 | 50,651 | 306,197 |
| | ND_NumIntentRet2 | Nondonor eggs, All patients 35-37 | 29,766 | |
| | ND_NumIntentRet3 | Nondonor eggs, All patients 38-40 | 28,917 | |
| | ND_NumIntentRet4 | Nondonor eggs, All patients 41-42 | 14,483 | |
| | ND_NumIntentRet5 | Nondonor eggs, All patients ≥43 | 11,725 | |

Due to a different reporting style of CDC data between 2014-2016 we have used SART data (**Table S2**) to estimate multiplication factor (**Table S3**) and consequently convert it to CDC format (**Table S4**) further used for forecasting of COVID-19. Here, as discussed in the manuscript a multiplication factor was used and the values of conversion are presented.

**Table S2.** Number of cycles between 2014-2018 for a patient's own eggs- live births per intended egg retrieval (all embryo transfers) extracted from https://www.sartcorsonline.com/Csr/Public.

| Year of record | Age group (years) | | | | |
|---|---|---|---|---|---|
| | < 35 | 35 - 37 | 38 - 40 | 41 - 42 | > 42 |
| 2014 | 41,063 | 21,407 | 20,732 | 11,106 | 8,611 |
| 2015 | 44,268 | 23,689 | 22,999 | 12,281 | 9,714 |
| 2016 | 46,189 | 25,448 | 24,495 | 12,601 | 9,784 |
| 2017 | 44,191 | 25,876 | 24,503 | 12,258 | 8,675 |
| 2018 | 42,820 | 26,027 | 24,957 | 12,686 | 8,931 |

**Table S3.** Multiplication factor per each age strata.

| Age of woman | < 35 | 35 - 37 | 38 - 40 | 41 - 42 | > 42 |
|---|---|---|---|---|---|
| Average multiplication factor | 1.18 | 1.13 | 1.16 | 1.16 | 1.33 |

**Table S4.** Outcomes of conversion SART to CDC.

| Year of record | Age group (years) | | | | |
|---|---|---|---|---|---|
| | < 35 | 35 - 37 | 38 - 40 | 41 - 42 | > 42 |
| 2018 | 50,651 | 29,766 | 28,917 | 14,483 | 11,725 |
| 2017 | 52,428 | 28,996 | 28,287 | 14,358 | 11,604 |
| 2016 | 54,747 | 28,812 | 28,335 | 14,575 | 12,968 |
| 2015 | 52,470 | 26,821 | 26,605 | 14,205 | 12,875 |
| 2014 | 48,671 | 24,237 | 23,982 | 12,846 | 11,413 |

**Table S5.** Total number of infants born, number of ART infants and the proportion of infants born from ART within the US for 1999 to 2018.

| Year | Nr. of infants born* | Nr. of ART infants† | Percentage of infants born from ART |
|---|---|---|---|
| 1999 | 3,959,417 | 30,967 | 0.78% |
| 2000 | 4,058,814 | 35,025 | 0.86% |
| 2001 | 4,025,933 | 40,687 | 1.01% |
| 2002 | 4,021,726 | 45,751 | 1.14% |
| 2003 | 4,089,950 | 48,756 | 1.19% |
| 2004 | 4,112,052 | 49,458 | 1.20% |
| 2005 | 4,138,349 | 52,041 | 1.26% |
| 2006 | 4,265,555 | 54,656 | 1.28% |
| 2007 | 4,316,233 | 57,569 | 1.33% |
| 2008 | 4,247,694 | 61,426 | 1.45% |
| 2009 | 4,130,665 | 60,190 | 1.46% |
| 2010 | 3,999,386 | 61,564 | 1.54% |
| 2011 | 3,953,590 | 61,610 | 1.56% |
| 2012 | 3,952,841 | 65,151 | 1.65% |
| 2013 | 3,932,181 | 66,691 | 1.70% |
| 2014 | 3,988,076 | 68,782 | 1.73% |
| 2015 | 3,978,497 | 71,152 | 1.79% |
| 2016 | 3,945,875 | 76,892 | 1.95% |
| 2017 | 3,855,500 | 78,052 | 2.02% |
| 2018 | 3,791,712 | 81,478 | 2.15% |

*Data obtained from U.S. Natality files (Birth Cohort dataset) compiled annually by the Center for Disease Control and Prevention's National Center for Health Statistics (NCHS).

†ART infants obtained from the annual CDC ART Success Rates Reports

For prediction of the number of cycles that would have occurred if the underlying growth had continued, we used the R 4.0.0 software environment with the "Forecast" package to perform auto regressive integrated moving average (ARIMA) prediction. This method assumes that future activity can be predicted based on past trends and no other variables. It also assumes a constant variance in errors around predictions, and no sudden changes in activity. We have therefore used it to predict clinical activity in 2009 to 2012 and 2020 to 2023 in the absence of external changes due to the 2008 recession and COVID-19 pandemic respectively.

**Table S6.** Prediction of total number of infants born, number of ART infants and the proportion of infants born from ART within the US for 2019 to 2023.

| Year | Predicted number | Lower 95% CI | Upper 95% CI |
|---|---|---|---|
| **5-year prediction of number of infants to be born** | | | |
| 2019 | 3,782,792 | 3,662,758 | 3,902,826 |
| 2020 | 3,777,939 | 3,538,256 | 4,017,623 |
| 2021 | 3,773,558 | 3,449,843 | 4,097,273 |
| 2022 | 3,769,231 | 3,378,530 | 4,159,931 |
| 2023 | 3,764,910 | 3,317,069 | 4,212,751 |
| **5-year prediction of number of ART infants to be born** | | | |
| 2019 | 84,134 | 80,395 | 87,873 |
| 2020 | 86,782 | 81,720 | 91,844 |
| 2021 | 89,424 | 83,476 | 95,371 |
| 2022 | 92,059 | 85,460 | 98,659 |
| 2023 | 94,690 | 87,588 | 101,792 |
| **5-year prediction of percentage of proportion of infants to be born** | | | |
| 2019 | 2.21 | 2.12 | 2.3 |
| 2020 | 2.27 | 2.15 | 2.4 |
| 2021 | 2.34 | 2.20 | 2.48 |
| 2022 | 2.40 | 2.25 | 2.55 |
| 2023 | 2.47 | 2.31 | 2.63 |

A quantitative prediction model was built using CDC data from 1999 to 2008, with four years onward prediction for 2009 to 2012 (Table S7). A similar quantitative model was built to predict age-stratified cycle starts for 2020 to 2023, using baseline data from 2014 to 2018. As a sensitivity analyses we also modelled only using 5 years data from 2004 to 2008 to predict 2009 to 2012 activity levels as 5 years data (2014 to 2018) was used to predict 2020 to 2023 (Table S8)

**Table S7.** Reported number of cycles and predicted number of cycles for 2009 to 2012.

| | **Reported Nr. of cycles †** | **Percentage reduction in cycles** | **Predicted number of cycles Nr. of cycles (95% CI)*** | | **Difference (95% CI)** | |
|---|---|---|---|---|---|---|
| | Year: 2009 | | | | | |
| **< 35** | 42,384 | -5.16% (-8.93~-1.07) | 44,692 | (42,842-46,542) | -2,308 | (-2,404~-2,212) |
| **35-37** | 21,860 | -9.00% (-14.51~-2.74) | 24,023 | (22,476-25,570) | -2,163 | (-2,302~-2,024) |
| **38-40** | 22,144 | -3.76% (-6.98~-0.31) | 23,009 | (22,212-23,806) | -865 | (-895~-835) |
| **41-42** | 9,845 | -2.85% (-6.31~0.87) | 10,134 | (9,760-10,508) | -289 | (-300~-278) |
| **Total** | **96,233** | **-5.52% (-7.84~-3.08)** | **101,858** | **(99,291-104,425)** | **-5,625** | **(-5,788~-5,462)** |
| | Year: 2010 | | | | | |
| **< 35** | 41,741 | -9.75% (-16.95~-1.16) | 46,248 | (42,232-50,263) | -4,507 | (-4,898~-4,115) |
| **35-37** | 21,366 | -14.06% (-21.57~-4.96) | 24,862 | (22,482-27,242) | -3,496 | (-3,831~-3,161) |
| **38-40** | 21,739 | -10.2% (-16.29~-3.16) | 24,208 | (22,448-25,969) | -2,469 | (-2,649~-2,290) |
| **41-42** | 10,120 | -4.11% (-11.10~4.07) | 10,554 | (9,724-11,383) | -434 | (-468~-399) |
| **Total** | **94,966** | **-10.3% (-14.39~-5.80)** | **105,872** | **(100,815-110,929)** | **-10,906** | **(-11,441~-10,371)** |
| | Year: 2011 | | | | | |
| **< 35** | 42,059 | -12.21% (-22.26~0.81) | 47,910 | (41,721-54,099) | -5,851 | (-6,607~-5,095) |
| **35-37** | 20,963 | -18.56% (-27.14~-7.68) | 25,740 | (22,707-28,772) | -4,777 | (-5,339~-4,214) |
| **38-40** | 21,128 | -16.81% (-24.84~-6.86) | 25,397 | (22,685-28,109) | -4,269 | (-4,725~-3,813) |
| **41-42** | 10,733 | -2.52% (-12.48~10.00) | 11,010 | (9,757-12,263) | -277 | (-309~-245) |
| **Total** | **94,883** | **-13.79% (-19.30~-7.47)** | **110,057** | **(102,545-117,569)** | **-15,174** | **(-16,189~-14,159)** |
| | Year: 2012 | | | | | |
| **< 35** | 41,798 | -15.8% (-27.82~1.01) | 49,644 | (41,381-57,907) | -7,846 | (-9,152~-6,540) |
| **35-37** | 20,920 | -21.43% (-30.74~-9.24) | 26,627 | (23,050-30,204) | -5,707 | (-6,474~-4,940) |
| **38-40** | 19,556 | -26.42% (-35.21~-14.87) | 26,578 | (22,971-30,185) | -7,022 | (-7,975~-6,069) |
| **41-42** | 10,740 | -6.49% (-18.13~8.99) | 11,486 | (9,854-13,119) | -746 | (-853~-640) |
| **Total** | **93,014** | **-18.65% (-25.09~-10.99)** | **114,335** | **(104,499-124,171)** | **-21,321** | **(-23,053~-19,589)** |

† number of fresh embryo nondonor cycles

*Prediction based on ARIMA modelling using data from 1999 to 2008 as baseline

**Table S8.** Reported number of cycles and predicted number of cycles for 2009 to 2012 using 5 years of baseline data for prediction.

| | Reported Nr. of cycles † | Percentage reduction in cycles | Predicted number of cycles Nr. of cycles (95% CI)* | | Difference (95% CI) | |
|---|---|---|---|---|---|---|
| | Year: 2009 | | | | | |
| **< 35** | 42,384 | -3.81% (-6.52~-0.93) | 44,062 | (42,783~45,341) | -1,678 | (-1,727~-1,629) |
| **35-37** | 21,860 | -5.75% (-11.8~1.19) | 23,193 | (21,602~24,785) | -1,333 | (-1,425~-1,242) |
| **38-40** | 22,144 | -2.79% (-6.63~1.38) | 22,779 | (21,843~23,716) | -635 | (-662~-609) |
| **41-42** | 9,845 | -2.67% (-5.76~0.63) | 10,115 | (9,783~10,447) | -270 | (-279~-261) |
| **Total** | **96,233** | **-3.91% (-6.04~-1.68)** | **100,149** | **(97,878~102,420)** | **-3,916** | **(-4,019~-3,813)** |
| | Year: 2010 | | | | | |
| **< 35** | 41,741 | -6.60% (-11.45~-1.19) | 44,691 | (42,244~47,138) | -2,950 | (-3,112~-2,788) |
| **35-37** | 21,366 | -8.53% (-22.51~11.61) | 23,358 | (19,144~27,572) | -1,992 | (-2,351~-1,633) |
| **38-40** | 21,739 | -7.74% (-15.52~1.62) | 23,563 | (21,392~25,734) | -1,824 | (-1,992~-1,656) |
| **41-42** | 10,120 | -3.27% (-9.68~4.13) | 10,462 | (9,719~11,205) | -342 | (-366~-318) |
| **Total** | **94,966** | **-6.96% (-11.63~-1.78)** | **102,074** | **(96,688~107,460)** | **-7,108** | **(-7,533~-6,683)** |
| | Year: 2011 | | | | | |
| **< 35** | 42,059 | -7.25% (-13.21~-0.43) | 45,349 | (42,239~48,458) | -3,290 | (-3,515~-3,064) |
| **35-37** | 20,963 | -11.61% (-31.42~24.30) | 23,717 | (16,865~30,568) | -2,754 | (-3,549~-1,958) |
| **38-40** | 21,128 | -13.02% (-22.92~-0.19) | 24,291 | (21,169~27,412) | -3,163 | (-3,569~-2,756) |
| **41-42** | 10,733 | -0.73% (-9.44~9.85) | 10,812 | (9,771~11,852) | -79 | (-86~-71) |
| **Total** | **94,883** | **-8.91% (-15.57~-1.12)** | **104,169** | **(95,957~112,381)** | **-9,286** | **(-10,189~-8,383)** |
| | Year: 2012 | | | | | |
| **< 35** | 41,798 | -9.13% (-15.85~-1.25) | 46,000 | (42,328~49,672) | -4,202 | (-4,537~-3,867) |
| **35-37** | 20,920 | -13.56% (-37.67~40.97) | 24,201 | (14,840~33,563) | -3,281 | (-4,551~-2,012) |
| **38-40** | 19,556 | -21.79% (-32.32~-7.37) | 25,003 | (21,112~28,893) | -5,447 | (-6,294~-4,599) |
| **41-42** | 10,740 | -3.78% (-13.66~8.65) | 11,162 | (9,885~12,439) | -422 | (-470~-374) |
| **Total** | **93,014** | **-12.55% (-20.65~-2.61)** | **106,366** | **(95,508~117,224)** | **-13,352** | **(-14,861~-11,843)** |

† number of fresh embryo nondonor cycles

*Prediction based on ARIMA modelling using data from 2004 to 2008 as baseline

**Table S9.** Predicted number of live-births after 2008 financial recession and observed number.

| | Reported Nr. of cycles † | Percentage of cycles resulting in live births (95% CI) * | Predicted number of cycles | Number of live births based on reported nr. of cycles | Estimated number of live births based on predicted nr. of cycles (95% CI) | Change from expected |
|---|---|---|---|---|---|---|
| | Year: 2009 | | | | | |
| **< 35** | 42,384 | 41.2% (40.73%, 41.67%) | 44,692 | 17,462 | 18,413 (18,203-18,623) | -951 (-940~-962) |
| **35-37** | 21,860 | 31.6% (30.99%, 32.22%) | 24,023 | 6,908 | 7,591 (7,443-7,739) | -683 (-669~-696) |
| **38-40** | 22,144 | 22.3% (21.76%, 22.85%) | 23,009 | 4,938 | 5,131 (5,006-5,256) | -193 (-187~-196) |
| **41-42** | 9,845 | 12.4% (11.76%, 13.07%) | 10,134 | 1,221 | 1,257 (1,191-1,323) | -36 (-33~-36) |
| **Total** | **96,233** | | **101,858** | **30,529** | **32,392 (32,099-32,685)** | **-1,863 (-1,846~-1,880)** |
| | Year: 2010 | | | | | |
| **< 35** | 41,741 | 41.5% (41.03%, 41.97%) | 46,248 | 17,323 | 19,193 (18,975-19,411) | -1,870 (-1,849~-1,892) |
| **35-37** | 21,366 | 31.9% (31.28%, 32.53%) | 24,862 | 6,816 | 7,931 (7,776-8,086) | -1,115 (-1,093~-1,136) |
| **38-40** | 21,739 | 22.1% (21.55%, 22.66%) | 24,208 | 4,804 | 5,350 (5,216-5,484) | -546 (-531~-558) |
| **41-42** | 10,120 | 12.4% (11.77%, 13.06%) | 10,554 | 1,255 | 1,309 (1,241-1,377) | -54 (-50~-55) |
| **Total** | **94,966** | | **105,872** | **30,198** | **33,783 (33,476-34,090)** | **-3,585 (-3,552~-3,618)** |
| | Year: 2011 | | | | | |
| **< 35** | 42,059 | 40% (39.53%, 40.47%) | 47,910 | 16,824 | 19,164 (18,939-19,389) | -2,340 (-2,313~-2,368) |
| **35-37** | 20,963 | 31.9% (31.27%, 32.53%) | 25,740 | 6,687 | 8,211 (8,049-8,373) | -1,524 (-1,494~-1,554) |
| **38-40** | 21,128 | 21.5% (20.95%, 22.06%) | 25,397 | 4,543 | 5,460 (5,319-5,601) | -917 (-893~-940) |
| **41-42** | 10,733 | 12.1% (11.5%, 12.73%) | 11,010 | 1,299 | 1,332 (1,264-1,400) | -33 (-30~-34) |
| **Total** | **94,883** | | **110,057** | **29,353** | **34,167 (33,849-34,485)** | **-4,814 (-4,769~-4,859)** |
| | Year: 2012 | | | | | |

| | | | | | | |
|---|---|---|---|---|---|---|
| **< 35** | 41,798 | 40.5% (40.03%, 40.97%) | 49,644 | 16,928 | 20,106 (19,873-20,339) | -3,178 (-3,141~-3,214) |
| **35-37** | 20,920 | 31.3% (30.68%, 31.93%) | 26,627 | 6,548 | 8,334 (8,167-8,501) | -1,786 (-1,749~-1,821) |
| **38-40** | 19,556 | 22.2% (21.62%, 22.79%) | 26,578 | 4,341 | 5,900 (5,744-6,056) | -1,559 (-1,516~-1,599) |
| **41-42** | 10,740 | 11.7% (11.11%, 12.32%) | 11,486 | 1,257 | 1,344 (1,275-1,413) | -87 (-82~-90) |
| **Total** | **93,014** | | **114,335** | **29,074** | **35,684 (35,350-36,018)** | **-6,610 (-6,546~-6,674)** |

† number of fresh embryo nondonor cycles
*The Confidence interval of a proportion

**Table S10.** Estimated changes in IVF within-cycle live-birth rate over the period 2020-2023 with implication of economic crisis triggered by COVID-19 by year.

| Year | Without Covid-19 impact Within-cycle live-birth rate (95% CI) | | With Covid-19 impact Within-cycle live-birth rate (95% CI) | | Difference (95% CI) | |
|---|---|---|---|---|---|---|
| **2020** | 34.0% | (33.7%~34.2%) | 33.3% | (33.0%~33.5%) | -0.7% | (-1.0%~-0.3%) |
| **2021** | 33.9% | (33.7%~34.1%) | 33.6% | (33.3%~33.5%) | -0.3% | (-0.7%~0.0%) |
| **2022** | 33.8% | (33.6%~34.1%) | 33.4% | (33.1%~33.6%) | -0.5% | (-0.8%~-0.1%) |
| **2023** | 33.8% | (33.5%~34.0%) | 33.5% | (33.2%~33.7%) | -0.3% | (-0.7%~0.1%) |

**Table S11.** Estimated IVF within-cycle live-birth rate over the period 2020-2023 with implication of 2 months treatment shutdown triggered by COVID-19 by age of patients.

| **Age group (years)** | **Within-cycle live-birth rate (95% CI)** | | | |
|---|---|---|---|---|
| | **Year: 2020** | | **Years: 2021-2023** | |
| **< 35** | 51.3% | (50.8%~51.7%) | 51.6% | (51.2%~52.0%) |
| **35-37** | 36.8% | (36.2%~37.3%) | 37.5% | (36.9%~38.1% |
| **38-40** | 22.9% | (22.4%~23.4%) | 23.5% | (23.0%~24.0%) |
| **41-42** | 11.1% | (10.6%~11.6%) | 11.8% | (11.3%~12.3% |
| **>42** | 3.2% | (2.9%~3.5%) | 3.4% | (3.1%~3.7%) |

### 2. Outcomes of COVID-19 with lower economic impact

**Table S12.** Estimated changes in number of IVF fresh nondonor cycles over the period 2020-2023 with implications of a less severe economic decline by reducing the annual effect by 50% from the estimations of economic crisis triggered by COVID-19 by age of patients.

| Age group (years) | Without COVID-19 impact Nr. of cycles (95% CI) | | Percentage reduction in cycles | With COVID-19 impact Nr. of cycles (95% CI) | | Difference (95% CI) | |
|---|---|---|---|---|---|---|---|
| | Year: 2020 | | | | | | |
| **< 35** | 51,170 | (43,798~58,542) | -2.58% | 49,850 | (42,668~57,032) | -1,320 | (-1,510~-1,130) |
| **35-37** | 31,358 | (28,389~34,327) | -4.50% | 29,947 | (27,112~32,782) | -1,411 | (-1,545~-1,277) |
| **38-40** | 30,385 | (27,296~33,474) | -1.88% | 29,814 | (26,783~32,845) | -571 | (-629~-513) |
| **41-42** | 14,924 | (13,234~16,615) | -1.43% | 14,711 | (13,045~16,377) | -213 | (-238~-189) |
| **>42** | 11,923 | (8,691~15,155) | -1.43% | 11,753 | (8,567~14,939) | -170 | (-216~-124) |
| **Total** | **139,760** | **(130,486~149,034)** | **-2.64%** | **136,075** | **(127,034~145,116)** | **-3,685** | **(-3,918~-3,452)** |
| | Year: 2021 | | | | | | |
| **< 35** | 51,835 | (36,545~67,126) | -4.88% | 49,305 | (34,761~63,849) | -2,530 | (-3,277~-1,784) |
| **35-37** | 32,771 | (27,096~38,446) | -7.03% | 30,467 | (25,191~35,743) | -2,304 | (-2,703~-1,905) |
| **38-40** | 31,704 | (25,583~37,825) | -5.10% | 30,087 | (24,278~35,896) | -1,617 | (-1,929~-1,305) |
| **41-42** | 15,469 | (11,670~19,268) | -2.06% | 15,150 | (11,429~18,871) | -319 | (-397~-241) |
| **>42** | 11,935 | (8,553~15,318) | -2.06% | 11,689 | (8,376~15,002) | -246 | (-316~-177) |
| **Total** | **143,714** | **(125,566~161,862)** | **-4.88%** | **136,698** | **(119,437~153,959)** | **-7,016** | **(-7,903~-6,129)** |
| | Year: 2022 | | | | | | |
| **< 35** | 52,499 | (32,207~72,792) | -6.11% | 49,291 | (30,238~68,344) | -3,208 | (-4,448~-1,969) |
| **35-37** | 34,222 | (27,014~41,431) | -9.28% | 31,046 | (24,506~37,586) | -3,176 | (-3,845~-2,508) |
| **38-40** | 33,041 | (25,126~40,956) | -8.40% | 30,266 | (23,016~37,516) | -2,775 | (-3,440~-2,110) |
| **41-42** | 16,034 | (10,707~21,362) | -1.26% | 15,832 | (10,572~21,092) | -202 | (-270~-135) |

| | | | | | | | |
|---|---|---|---|---|---|---|---|
| **>42** | 11,913 | (8,507~15,318) | -1.26% | 11,763 | (8,400~15,126) | -150 | (-192~-107) |
| **Total** | **147,709** | **(123,910~171,508)** | **-6.44%** | **138,198** | (115,898~160,498) | **-9,511** | (-11,009~-8,013) |
| | Year: 2023 | | | | | | |
| **< 35** | 53,163 | (28,878~77,448) | -7.90% | 48,963 | (26,597~71329) | -4,200 | (-6,119~-2,281) |
| **35-37** | 35,665 | (27,153~44,177) | -10.71% | 31,845 | (24,245~39445) | -3,820 | (-4,732~-2,908) |
| **38-40** | 34,376 | (24,987~43,765) | -13.21% | 29,835 | (21,686~37984) | -4,541 | (-5,781~-3,301) |
| **41-42** | 16,603 | (10,059~23,147) | -3.24% | 16,065 | (9,733~22397) | -538 | (-750~-326) |
| **>42** | 11,883 | (8,474~15,293) | -3.24% | 11,498 | (8,199~14797) | -385 | (-496~-275) |
| **Total** | **151,690** | **(123,321~180,059)** | **-8.89%** | **138,206** | **(112,218~164194)** | **-13,484** | **(-15,865~-11,103)** |

**Table S13.** Estimated changes in number of IVF live births over the period 2020-2023 with implication of lower economic impact triggered by COVID-19 by age of patients.

| Age group (years) | Without COVID-19 impact Live births (95% CI) | | COVID-19 with lower economic impact. Live births (95% CI) | | Difference (95% CI) | |
|---|---|---|---|---|---|---|
| | Year: 2020 | | | | | |
| **< 35** | 26,404 | (22,593~30,214) | 25,561 | (21,872-29,250) | -843 | (-941~-745) |
| **35-37** | 11,759 | (10,632~12,886) | 11,006 | (9,951-12,061) | -753 | (-803~-703) |
| **38-40** | 7,140 | (6,399~7,882) | 6,836 | (6,126-7,546) | -305 | (-318~-291) |
| **41-42** | 1,761 | (1,547~1,975) | 1,634 | (1,435-1,833) | -127 | (-130~-124) |
| **>42** | 405 | (289~522) | 374 | (266-482) | -31 | (-33~-30) |
| **Total** | **47,470** | **(43,421~51,519)** | **45,411** | **(41,502-49,319)** | **-2,059** | **(-2,170~-1,948)** |
| | Year: 2021 | | | | | |
| **< 35** | 26,747 | (18,854~34,640) | 25,441 | (17,934-32,949) | -1,305 | (-1,691~-920) |
| **35-37** | 12,289 | (10,153~14,425) | 11,425 | (9,439-13,411) | -864 | (-1,014~-714) |
| **38-40** | 7,450 | (6,003~8,897) | 7,070 | (5,697-8,444) | -380 | (-454~-306) |
| **41-42** | 1,825 | (1,370~2,281) | 1,788 | (1,341-2,234) | -38 | (-47~-28) |
| **>42** | 406 | (284~527) | 397 | (278-516) | -8 | (-11~-6) |
| **Total** | **48,718** | **(40,400~57,035)** | **46,122** | **(38,222-54,022)** | **-2,595** | **(-3,016~-2,175)** |
| | Year: 2022 | | | | | |
| **< 35** | 27,089 | (16,616~37,563) | 25,434 | (15,601-35,268) | -1,655 | (-2,295~-1,015) |
| **35-37** | 12,833 | (10,123~15,543) | 11,642 | (9,184-14,101) | -1,191 | (-1,442~-940) |
| **38-40** | 7,765 | (5,897~9,632) | 7,113 | (5,402-8,823) | -652 | (-809~-495) |
| **41-42** | 1,892 | (1,258~2,526) | 1,868 | (1,242-2,494) | -24 | (-32~-16) |
| **>42** | 405 | (283~527) | 400 | (279-521) | -5 | (-7~-4) |
| **Total** | **49,984** | **(38,987~60,982)** | **46,457** | **(36,158-56,756)** | **-3,527** | **(-4,233~-2,822)** |
| | Year: 2023 | | | | | |
| **< 35** | 27,432 | (14,899~39,965) | 13,722 | (13,722-36,808) | -2,167 | (-3,157~-1,177) |
| **35-37** | 13,374 | (10,176~16,573) | 9,086 | (9,086-14,798) | -1,433 | (-1,775~-1,090) |
| **38-40** | 8,078 | (5,865~10,291) | 5,091 | (5,091-8,932) | -1,067 | (-1,359~-775) |
| **41-42** | 1,959 | (1,182~2,736) | 1,144 | (1,144-2,648) | -63 | (-89~-38) |
| **>42** | 404 | (282~526) | 273 | (273-509) | -13 | (-17~-9) |
| **Total** | **51,248** | **(38,102~64,394)** | **34,436** | **(34,436-58,574)** | **-4,743** | **(-5,831~-3,655)** |

#### 3. Outcomes of COVID-19 with greater economic impact

**Table S14.** Estimated changes in number of IVF fresh nondonor cycles over the period 2020-2023 with implication of more severe decline by increasing it by 50% of the estimations of economic crisis triggered by COVID-19 by age of patients.

| Age group (years) | Without COVID-19 impact Nr. of cycles (95% CI) | | Percentage reduction in cycles | With COVID-19 impact Nr. of cycles (95% CI) | | Difference (95% CI) | |
|---|---|---|---|---|---|---|---|
| | Year: 2020 | | | | | | |
| **< 35** | 51,170 | (43,798~58,542) | -7.74% | 47,209 | (40,408~54,010) | 3,961 | (-4,532~-3,390) |
| **35-37** | 31,358 | (28,389~34,327) | -13.5% | 27,125 | (24,557~29,693) | -4,233 | (-4,634~-3,832) |
| **38-40** | 30,385 | (27,296~33,474) | -5.64% | 28,671 | (25,756~31,586) | -1,714 | (-1,888~-1,540) |
| **41-42** | 14,924 | (13,234~16,615) | -4.28% | 14,285 | (12,667~15,903) | -639 | (-712~-567) |
| **>42** | 11,923 | (8,691~15,155) | -4.28% | 11,413 | (8,319~14,507) | -510 | (-648~-372) |
| **Total** | **139,760** | **(130,486~149,034)** | **-7.91%** | **128,703** | **(120,127~137,279)** | **-11,057** | **(-11,755~-10,359)** |
| | Year: 2021 | | | | | | |
| **< 35** | 51,835 | (36,545~67,126) | -14.63% | 44,252 | (31,198~57,306) | -7,583 | (-9,820~-5,347) |
| **35-37** | 32,771 | (27,096~38,446) | -21.09% | 25,860 | (21,382~30,338) | -6,911 | (-8,108~-5,714) |
| **38-40** | 31,704 | (25,583~37,825) | -15.30% | 26,853 | (21,669~32,037) | -4,851 | (-5,789~-3,914) |
| **41-42** | 15,469 | (11,670~19,268) | -6.17% | 14,515 | (10,950~18,080) | -954 | (-1,188~-720) |
| **>42** | 11,935 | (8,553~15,318) | -6.17% | 11,199 | (8,025~14,373) | -736 | (-945~-528) |
| **Total** | **143,714** | **(125,566~161,862)** | **-14.64%** | **122,679** | **(107,183~138,175)** | **-21,035** | **(-23,687~-18,383)** |
| | Year: 2022 | | | | | | |
| **< 35** | 52,499 | (32,207~72,792) | -18.32% | 42,881 | (26,306~59,456) | -9,618 | (-13,336~-5,901) |
| **35-37** | 34,222 | (27,014~41,431) | -27.84% | 24,695 | (19,493~29,897) | -9,527 | (-11,534~-7,521) |
| **38-40** | 33,041 | (25,126~40,956) | -25.21% | 24,711 | (18,791~30,631) | -8,330 | (-10,325~-6,335) |
| **41-42** | 16,034 | (10,707~21,362) | -3.78% | 15,428 | (10,302~20,554) | -606 | (-808~-405) |

| | | | | | | | |
|---|---|---|---|---|---|---|---|
| **>42** | 11,913 | (8,507~15,318) | -3.78% | 11,463 | (8,186~14,740) | -450 | (-578~-321) |
| **Total** | **147,709** | **(123,910~171,508)** | **19.32%** | **119,178** | **(99,843~138,513)** | **-28,531** | **(-32995~-24067)** |
| | Year: 2023 | | | | | | |
| **< 35** | 53,163 | (28,878~77,448) | -23.70% | 40,563 | (22,034~59,092) | -12,600 | (-18,356~-6,844) |
| **35-37** | 35,665 | (27,153~44,177) | -32.14% | 24,202 | (18,426~29,978) | -11,463 | (-14,199~-8,727) |
| **38-40** | 34,376 | (24,987~43,765) | -39.63% | 20,753 | (15,085~26,421) | -13,623 | (-17,344~-9,902) |
| **41-42** | 16,603 | (10,059~23,147) | -9.73% | 14,988 | (9,081~20,895) | -1,615 | (-2,252~-978) |
| **>42** | 11,883 | (8,474~15,293) | -9.73% | 10,727 | (7,649~13,805) | -1,156 | (-1,488~-825) |
| **Total** | **151,690** | **(99,551~203,830)** | **26.67%** | **111,233** | **(89,945~132,521)** | **-40,457** | **(-47,538~-33,376)** |

**Table S15.** Estimated changes in number of IVF live births over the period 2020-2023 with implication of greater economic impact triggered by COVID-19 by age of patients.

| **Age group (years)** | **Without COVID-19 impact Live births (95% CI)** | | **COVID-19 with greater economic impact. Live births (95% CI)** | | **Difference (95% CI)** | |
|---|---|---|---|---|---|---|
| | Year: 2020 | | | | | |
| **< 35** | 26,404 | (22,593~30,214) | 24,207 | (20,713-27,700) | -2,197 | (-2,491~-1,903) |
| **35-37** | 11,759 | (10,632~12,886) | 9,969 | (9,013-10,925) | -1,790 | (-1,940~-1,641) |
| **38-40** | 7,140 | (6,399~7,882) | 6,574 | (5,891-7,257) | -567 | (-608~-526) |
| **41-42** | 1,761 | (1,547~1,975) | 1,587 | (1,393-1,780) | -174 | (-183~-165) |
| **>42** | 405 | (289~522) | 363 | (259-468) | -42 | (-47~-37) |
| **Total** | **47,470** | **(43,421~51,519)** | 42,699 | **(39,007-46,391)** | **-4,770** | **(-5103~-4,438)** |
| | Year: 2021 | | | | | |
| **< 35** | 26,747 | (18,854~34,640) | 22,834 | (16,096-29,572) | -3,913 | (-5,068~-2,758) |
| **35-37** | 12,289 | (10,153~14,425) | 9,698 | (8,012-11,383) | -2,592 | (-3,042~-2,141) |
| **38-40** | 7,450 | (6,003~8,897) | 6,310 | (5,085-7,536) | -1,140 | (-1,361~-919) |
| **41-42** | 1,825 | (1,370~2,281) | 1,713 | (1,285-2,140) | -113 | (-141~-84) |
| **>42** | 406 | (284~527) | 381 | (267-495) | -25 | (-33~-18) |
| **Total** | **48,718** | **(40,400~57,035)** | 40,936 | **(33,868-48,003)** | **-7,782** | **(-9,041~-6,523)** |
| | Year: 2022 | | | | | |
| **< 35** | 27,089 | (16,616~37,563) | 22,127 | (13,572-30,681) | -4,963 | (-6,882~-3,044) |
| **35-37** | 12,833 | (10,123~15,543) | 9,261 | (7,305-11,216) | -3,573 | (-4,327~-2,818) |
| **38-40** | 7,765 | (5,897~9,632) | 5,807 | (4,411-7,204) | -1,958 | (-2,428~-1,487) |
| **41-42** | 1,892 | (1,258~2,526) | 1,821 | (1,210-2,431) | -72 | (-95~-48) |
| **>42** | 405 | (283~527) | 390 | (272-507) | -15 | (-20~-11) |
| **Total** | **49,984** | **(38,987~60,982)** | **39,405** | **(30,497-48,312)** | **-10,580** | **(-12,695~-8,465)** |
| | Year: 2023 | | | | | |
| **< 35** | 27,432 | (14,899~39,965) | 20,931 | (11,368-30,493) | -6,502 | (-9,472~-3,531) |
| **35-37** | 13,374 | (10,176~16,573) | 9,076 | (6,905-11,246) | -4,299 | (-5,327~-3,271) |
| **38-40** | 8,078 | (5,865~10,291) | 4,877 | (3,541-6,213) | -3,201 | (-4,078~-2,324) |
| **41-42** | 1,959 | (1,182~2,736) | 1,769 | (1,067-2,470) | -191 | (-266~-115) |
| **>42** | 404 | (282~526) | 365 | (254-475) | -39 | (-51~-27) |
| **Total** | **51,248** | **(38,102~64,394)** | **37,017** | **(27,095-46,938)** | **-14,232** | **(-17,496~-10,967)** |
